# Ultra-processed foods and hypertension incidence in RaNCD cohort project

**DOI:** 10.1101/2024.06.13.24308899

**Authors:** Parsa Amirian, Mahsa Zarpoosh, Yahya Pasdar

## Abstract

**Background:** Due to rapid population growth and, subsequently, large-scale food production methods, ultra-processed food consumption is in parabolic growth. By affecting 1.28 billion adults globally, hypertension is a major risk factor and cause of premature death. In order to find the relation between ultra-processed food consumption and other covariates with hypertension incidence, this study was conducted in the western part of Iran using RaNCD prospective cohort data.

**Methods:** We included 8150 participants at the risk of hypertension in the final analysis. Using the data obtained from the Iranian food frequency questionnaire and the NOVA food classification, we assessed the ultra-processed food consumption of each participant in a day. Logistic regression models and the Cox proportional hazards regression model were used to assess the association between ultra-processed food consumption and hypertension in the main model and sensitivity analysis.

**Results:** The mean age of participants was 46.25y ± 7.94 (47.58% males); the mean follow-up time was 7.65y ± 1.62, and the mean daily UPF intake in g/d among participants was 88.07 ± 84.96. During the follow-up period, 862 cases of hypertension were recorded. We adjusted the main model for several confounders, including age, gender, residence type, marital status, socioeconomic status, physical activity, body mass index, familial history of hypertension, fasting blood sugar, and waist-to-hip ratio. The odds ratio and 95% confidence interval (95% CI) of the second and third tertile of UPFs were 1.15 (95% CI, 0.96-1.37) and 1.03 (95%CI, 0.85-1.24), respectively, compared to the first tertile with insignificant p-value & p-trend.

**Conclusion:** To the best of our knowledge, our study is the first to assess the association between hypertension and ultra-processed foods in the Middle East region. Significant associations between hypertension incidence and some confounders were also identified.

## Introduction

Since the introduction of NOVA food classification that separates foods and beverages into four categories, including unprocessed, processed culinary ingredients, processed, and ultra-processed foods (UPFs), the light was shed on an overlooked aspect of nutrition which is the journey that food takes from agricultural steps to when it is eaten (1). Rural-to-urban migration, agrarian transformation, population growth, heavy UFP marketing, and economic progress, alongside many other factors, led to excessive UPF consumption percentages in the population diet; for example, population growth and the demand for food worldwide led the economy to produce food in large scales which are mostly UPFs, therefore understanding the effects of UPF consumption in many aspects of health is crucial (2). Being a major risk factor and health concern, as said by the World Health Organization (WHO), 1.28 billion adults suffer from hypertension (HTN); it is a substantial reason for premature death around the globe (3). The HTN prevalence is not equal in different parts of the world; low and middle-income countries have a higher prevalence than developed countries, which makes HTN-related research in regions like the Middle East and North Africa much more critical (3). To the best of our knowledge, the association between UPF consumption and HTN incidence, despite being a significant concern and area of interest, has been rarely investigated before, especially in low and middle-income countries. The UPF/HTN association was investigated before in countries like Spain and Brazil in a prospective cohort style with mixed results, Mendonça et al. used the Spanish Navarra Project data and found that participants in the highest tertile of UPF consumption had 21% higher chance of developing HTN compared to the first tertile (adjusted hazard ratio, 1.21; 95% CI, 1.06, 1.37); by using Poisson regression models Rezende-Alves et al. found that participants in the highest quintile of UPF energetic intake had the relative risk of 1.35 (95% CI, 1.01, 1.81) (4,5). Due to the high prevalence of HTN and different methods of processing food in different countries, it is crucial to assess the association of UPF/HTN in various populations, especially in the Middle East. The Ravansar non-communicable disease (RaNCD) cohort is the first cohort on the Iranian/Kurdish population in the western part of Iran; to obtain data regarding nutritional status, RaNCD uses a 113-item Food Frequency Questionnaire (FFQ); the main phase of the cohort was started in March 2015 with six completed follow-ups (6). This study was conducted to fill the gap in our knowledge regarding UPF/HTN relations in the Middle East region.

## Methods

### Study Population

We were provided with data from 10047 participants registered in the RaNCD prospective cohort, aged over 35; all participants resided in Ravansar County in the northwestern part of Kermanshah province (6). Firstly, we overviewed the overall data; we excluded 192 participants due to missing data; furthermore, 1579 participants had already been diagnosed with HTN or were using HTN drugs. Participants with improbable energy intake > 5500 kcal (n = 102) and < 800 kcal (n = 24) were also excluded. Finally, 8150 participants were incorporated in the final analysis (Figure 1).

**Figure 1.**
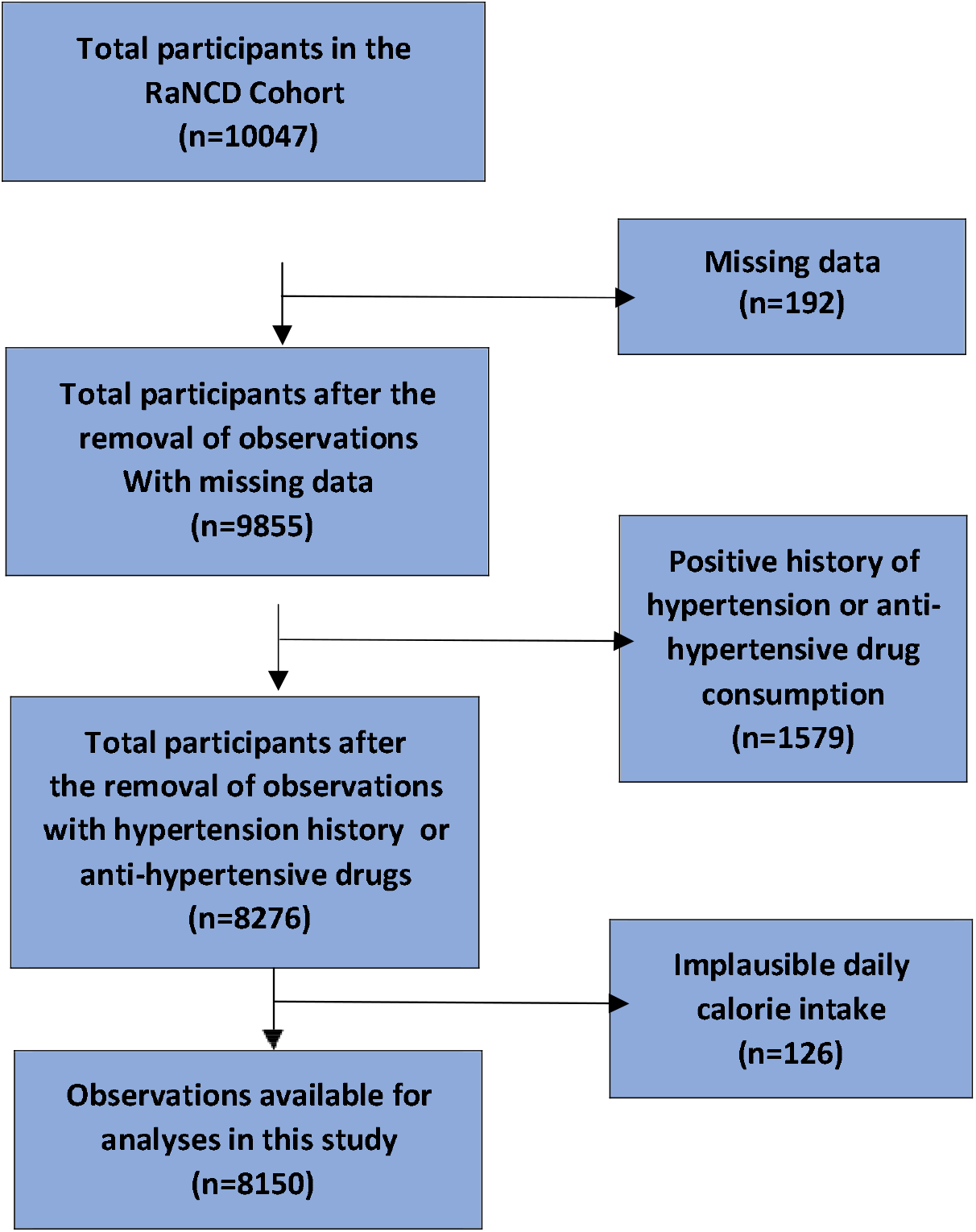
Flow-chart of participants.

### Nutritional Status

Using the Iranian Food Frequency Questionnaire (FFQ), 113 food groups and an additional four local foods were handed over to us for the investigation. We used the NOVA food classification system and designated each food group into one of the four NOVA groups (unprocessed, processed culinary ingredients, processed, and UPFs). The results were as follows: 66 food groups were classified as unprocessed, 12 as processed culinary ingredients, 24 as processed, and 19 as UPFs. We found 19 UPFs as follows: 1. Baguette bread 2. Sausage/ salami 3. Hamburger 4. Pizza 5. Flavored milk 6. Margarine 7. Hydrogenated oil 8. Mayonnaise 9. Rock candy/ other sweets 10. Soda drinks 11. Nonalcoholic malt beverages 12. Ice cream 13. Dried cookies 14. Creamed cookies 15. Chocolate 16. Chips 17. Cheese puffs 18. Concentrated juice 19. Crackers/ biscuits (Table 1). All measurements were documented as gram/day (g/d).

**Table 1.**
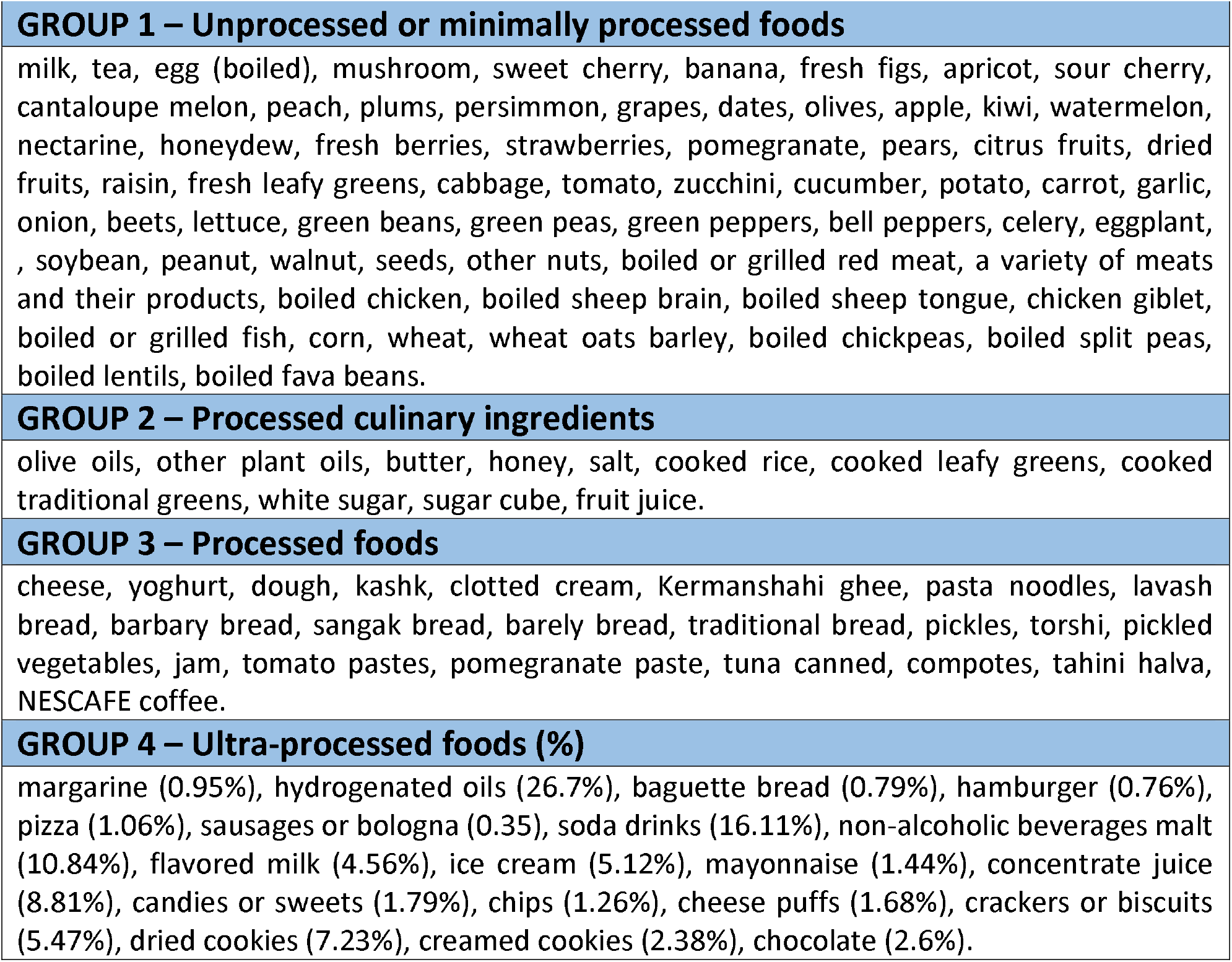
NOVA classification of PERSIAN cohort FFQ items.

|  |
| --- |
| **GROUP 1 – Unprocessed or minimally processed foods** |
| milk, tea, egg (boiled), mushroom, sweet cherry, banana, fresh figs, apricot, sour cherry, cantaloupe melon, peach, plums, persimmon, grapes, dates, olives, apple, kiwi, watermelon, nectarine, honeydew, fresh berries, strawberries, pomegranate, pears, citrus fruits, dried fruits, raisin, fresh leafy greens, cabbage, tomato, zucchini, cucumber, potato, carrot, garlic, onion, beets, lettuce, green beans, green peas, green peppers, bell peppers, celery, eggplant, , soybean, peanut, walnut, seeds, other nuts, boiled or grilled red meat, a variety of meats and their products, boiled chicken, boiled sheep brain, boiled sheep tongue, chicken giblet, boiled or grilled fish, corn, wheat, wheat oats barley, boiled chickpeas, boiled split peas, boiled lentils, boiled fava beans. |
| **GROUP 2 – Processed culinary ingredients** |
| olive oils, other plant oils, butter, honey, salt, cooked rice, cooked leafy greens, cooked traditional greens, white sugar, sugar cube, fruit juice. |
| **GROUP 3 – Processed foods** |
| cheese, yoghurt, dough, kashk, clotted cream, Kermanshahi ghee, pasta noodles, lavash bread, barbery bread, sangak bread, barely bread, traditional bread, pickles, torshi, pickled vegetables, jam, tomato pastes, pomegranate paste, tuna canned, compotes, tahini halva, NESCAFE coffee. |
| **GROUP 4 – Ultra-processed foods (%)** |
| margarine (0.95%), hydrogenated oils (26.7%), baguette bread (0.79%), hamburger (0.76%), pizza (1.06%), sausages or bologna (0.35), soda drinks (16.11%), non-alcoholic beverages malt (10.84%), flavored milk (4.56%), ice cream (5.12%), mayonnaise (1.44%), concentrate juice (8.81%), candies or sweets (1.79%), chips (1.26%), cheese puffs (1.68%), crackers or biscuits (5.47%), dried cookies (7.23%), creamed cookies (2.38%), chocolate (2.6%). |

### Hypertension Incidence

As previously mentioned, in the initial phase of the RaNCD cohort, 1579 were diagnosed with HTN or were using HTN-related drugs. In the follow-up phases, 1029 participants were identified to have HTN; after cleaning the data, we were left with 862 HTN incidences. HTN was defined by systolic blood pressure (SBP) ^3^ 140 mmHg and diastolic blood pressure (DBP) ^3^ 90 mmHg in two or more readings by the cohort crew or other physicians were considered as new incidences of HTN. The time at risk of HTN for included participants was 22,774,803 days.

### Statistical Analysis

We checked for several covariates including age, sex, residence types, marital status, body mass index (BMI), waist-to-hip ratio (WHR), socioeconomic status quartiles (SESq), history of fasting blood sugar (FBS), familial history of different diseases, metabolic equivalent (MET) quartiles, UPF tertiles, past medical history of different diseases, alcohol use, smoking status, total energy intake (Kcal/day), total carbohydrate intake (g/d), total lipid intake (g/d), total protein intake (g/d). By adding up values of each UPF item (g/d), we estimated the total grams of UPF consumed in a day; then, we created tertiles of UPFs (tertile one had the least consumption, and tertile three had the highest amount). Logistic regression models were used to analyze data and estimate odds ratios (ORs) and 95% confidence intervals (CIs) in the primary model and sensitivity analysis; furthermore, to catch temporal aspects of HTN incidence, we ran the main model using the Cox proportional hazards model. Potential confounders in the main model were gender, age, residence type, marital status, BMI, WHR, SESq, FBS, FH of HTN, and MET quartiles.

To assess the robustness of our model, we designed several scenarios in the context of sensitivity analysis. First, we introduced alcohol consumption and smoking status to our model as further adjustments; in the next model, we further adjusted the model for total energy, carbohydrate, lipid, and protein intake in a day; in the following scenario, we brought in FHs to the model namely familial histories of diabetes, cardiac diseases, myocardial infarction (MI), and stroke. In the next scenario, we excluded early cases (< 3 years) of HTN and executed the main model; next, we adjusted the main model for medical conditions, including Rheumatoid arthritis (RA), osteoporosis, muscle weakness, and weight loss. Subsequently, we checked for interactions between covariates, e.g., gender & age, gender & BMI, gender & UPF tertiles, BMI & UPF tertiles, and age & UPF tertiles; subgroup analysis was also performed; this analysis was based on age (<50 & >50), gender (male & female), residence type (urban & rural), FH of HTN (negative & positive).

## Results

### Participant Characteristics

We have included 8150 participants in our study (47.58% male, 52.42% female), the mean age ± SD was 46.25y ± 7.94, the mean follow-up time for HTN was 7.65y ± 1.62, and the mean daily UPF intake in g/d among participants was 88.07 ± 84.96; approximately, half of participants had a positive history of HTN (47.82%), and the mean BMI was 27.32± 4.61; Table 2 shows characteristics of participants in each UPF terile. RaNCD Participants in the third tertile of UPF consumption are more likely to be men, younger, live in the city, be smokers, have better SES, consume more energy (fourth quantile of energy intake), have almost the same physical activity and BMI compared to the first terile.

**Table 2.** Characteristics of participants in each UPF terile.

| Characteristics | Tertile 1 (N = 2735) | Tertile (N = 2746) | Tertile 3 (N = 2669) |
| --- | --- | --- | --- |
| | Mean ± SD or N (percentage) | | |
| UPFs intake (g/d) | 24.53 ± 11.58 | 63.83 ± 13.80 | 178.13 ± 94.11 |
| Age (years) | 47.87 ± 8.08 | 46.27 ± 7.93 | 44.58 ± 7.46 |
| Male | 969 (35.42) | 1274 (46.39) | 1635 (61.25) |
| Marital status |  |  |  |
| Single | 143 (5.22) | 141 (5.13) | 100 (3.74) |
| Residence type |  |  |  |
| Urban | 1425 (52.10) | 1536 (55.93) | 1891 (70.85) |
| Alcohol use |  |  |  |
| No | 2689 (98.31) | 2627 (95.66) | 2438(91.34) |
| BMI | 27.49 ± 4.61 | 27.11 ± 4.56 | 27.36 ± 4.65 |
| Smoking |  |  |  |
| No | 1253 (45.81) | 1177 (42.86) | 936 (35.06) |
| Current | 222 (8.11) | 312 (11.36) | 442 (16.56) |
| Former | 202 (7.38) | 222 (8.08) | 206 (7.71) |
| Passive | 1058 (38.68) | 1035 (37.69) | 1085 (40.65) |
| SES quantiles |  |  |  |
| 1 | 682 (24.94) | 572 (20.83) | 315 (11.80) |
| 2 | 550 (20.11) | 566 (20.61) | 482 (18.06) |
| 3 | 532 (19.45) | 518 (18.86) | 602 (22.56) |
| 4 | 479 (17.51) | 542 (19.74) | 621 (23.27) |
| 5 | 492 (17.99) | 548 (19.96) | 649 (24.32) |
| T2DM |  |  |  |
| Yes | 209 (48.37) | 125 (28.93) | 98 (22.68) |
| CVDs |  |  |  |
| YES | 232 (8.48) | 170 (6.19) | 124 (4.65) |
| MET |  |  |  |
| 1 | 650 (23.77) | 703 (25.60) | 700 (26.23) |
| 2 | 770 (28.15) | 657 (23.93) | 610 (22.86) |
| 3 | 760 (27.79) | 689 (25.09) | 613 (22.97) |
| 4 | 555 (20.29) | 697 (25.38) | 746 (27.95) |
| HTN (FH) |  |  |  |
| Yes | 1280 (46.80) | 1313 (47.82) | 1304 (48.86) |
| Energy (kcal) | 2224.68 ± 723.68 | 2620.77 ± 760.52 | 3248.80 ± 867.31 |

### Main model

A logistic regression model was used to fit the data as the primary model and to analyze the sensitivity of our main model; logistic regression models with different adjustments were used; to capture temporal aspects of HTN incidence, we used the Cox proportional hazard model as a sensitivity analysis; in the Cox model, the proportional hazards assumption showed no violation. The odds ratio (OR) & 95% confidence interval (95% CI) of the second and third tertile of UPFs were 1.15 (95% CI, 0.96-1.37) and 1.03 (95%CI, 0.85-1.24) respectively compared to the first tertile with insignificant p-value & p-trend (> 0.05). Alternatively, our main model showed significant associations as well; females had 31% higher chance of HTN development compared males, OR (95% CI, p-value) 1.31 (1.09–1.58, 0.003), in our population increasing the age by one year corresponds with 6% higher chance of HTN development, OR (95% CI, p-value) 1.06 (1.04-1.07, 0.000); the OR (95% CI, p-value) for HTN development in spouses and widows compared to singles were as follows 2.22 (1.25-3.94, 0.006) & 2.41 (1.26-4.59, 0.007); 1 unit increase in BMI corresponds to 4% chance of HTN development OR (95% CI, p-value) 1.04 (1.02-1.06, 0.000); surprisingly, the OR (95% CI, p-value) for WHR was 8.62 (1.82-40.92, 0.007); consequently, OR (95% CI, p-value) for FBS and familial history of HTN were as follows 1.002 (1.000-1.004, 0.028), 1.28 (1.11-1.49, 0.001) respectively.

### Sensitivity analysis

Several scenarios as sensitivity analyses were designed to test our main model’s robustness. Initially, we further adjusted the main model; these adjustments were as follows alcohol use, smoking status, total lipid consumption, total carbohydrate consumption, total protein consumption, total energy intake, familial histories of NCDs (type 2 diabetes, cardiac diseases, MI, and stroke), and PMHs (rheumatoid arthritis (RA), osteoporosis, gastroesophageal reflux disease (GERD); none showed significance except for familial history of stroke the OR (95% CI, p-value) was 1.31 (1.07-1.60, 0.008). Consequently, we excluded early cases of HTN incidence (<3y), and the results did not change dramatically compared to the main model (the p-value of FBS became insignificant). In the following scenario, we checked for interactions; the interaction between gender and BMI showed OR of 0.94 (0.91-0.98, 0.002), meaning that the impact of BMI on HTN incidence in females is approximately 6% less than males; other interactions were insignificant (p-value > 0.05). The results of the subgroup analysis can be found in Table 3. As previously mentioned, the Cox proportional hazard model was used to assess the temporal aspects of our main model. The results, including hazard ratios (HRs), p-values, and 95% CIs, can be found in Table 4.

**Table 3.** ORs and p-values of age, BMI, familial history of hypertension, WHR, and each UPF tertiles in the subgroup analysis.

| Subgroups | UPF tertiles |  |  | WHR | Age | BMI | FH HTN |
| --- | --- | --- | --- | --- | --- | --- | --- |
|  | Tertile 1 | Tertile 2 | Tertile 3 |  |  |  |  |

|  | OR | p-value | OR | p-value | OR | p-value | OR | p-value | OR | p-value | OR | p-value | HR | p-value |
| --- | --- | --- | --- | --- | --- | --- | --- | --- | --- | --- | --- | --- | --- | --- |
| Age > 50 | 1.00 | - | 1.10 | 0.434 | 0.94 | 0.772 | 17.2 | 0.022 | 1.02 | 0.153 | 1.04 | 0.024 | 0.97 | 0.812 |
| Age < 50 | 1.00 | - | 1.16 | 0.238 | 1.09 | 0.500 | 6.75 | 0.077 | 1.11 | 0.000 | 1.04 | 0.002 | 1.48 | 0.000 |
| Males | 1.00 | - | 1.10 | 0.517 | 0.84 | 0.290 | 0.50 | 0.611 | 1.07 | 0.000 | 1.12 | 0.000 | 1.46 | 0.002 |
| Females | 1.00 | - | 1.16 | 0.169 | 1.19 | 0.151 | 19.6 | 0.006 | 1.04 | 0.000 | 1.02 | 0.058 | 1.20 | 0.056 |
| Urban | 1.00 | - | 1.05 | 0.644 | 0.94 | 0.623 | 11.0 | 0.015 | 1.05 | 0.000 | 1.04 | 0.001 | 1.23 | 0.027 |
| Rural | 1.00 | - | 1.29 | 0.058 | 1.20 | 0.281 | 5.41 | 0.209 | 1.06 | 0.000 | 1.04 | 0.008 | 1.38 | 0.008 |
| FH HTN |  |  |  |  |  |  |  |  |  |  |  |  |  |  |
| Yes | 1.00 | - | 1.38 | 0.010 | 1.12 | 0.393 | 2.48 | 0.417 | 1.05 | 0.000 | 1.05 | 0.001 | - | - |
| No | 1.00 | - | 0.96 | 0.759 | 0.95 | 0.757 | 26.5 | 0.004 | 1.06 | 0.000 | 1.04 | 0.008 | - | - |

**Table 4.** HRs, p-values, and 95%CIs derived from Cox proportional hazard model.

| Variables | HR | p-value | 95%CI |
| --- | --- | --- | --- |
| Gender |  |  |  |
| Male | 1.00(ref) | - | - |
| Female | 1.28 | 0.004 | 1.08-1.52 |
| Age | 1.05 | 0.000 | 1.04-1.06 |
| UPF |  |  |  |
| Tertile 1 | 1.00(ref) | - | - |
| Tertile 2 | 1.13 | 0.116 | 0.96-1.33 |
| Tertile 3 | 1.01 | 0.881 | 0.85-1.20 |
| Marital status |  |  |  |
| single | 1.00(ref) | - | - |
| Married | 2.14 | 0.007 | 1.22-3.755 |
| Widowed | 2.30 | 0.008 | 1.24-4.26 |
| BMI | 1.04 | 0.000 | 1.02-1.06 |
| WHR | 7.54 | 0.004 | 1.89-30.1 |
| FBS | 1.002 | 0.017 | 1.000-1.004 |
| FH HTN | 1.24 | 0.002 | 1.08-1.42 |

## Discussion

To the best of our knowledge, our study, on the one hand, is the first cohort on the Iranian/Kurdish population (6); on the other hand, the first to assess the HTN/UPFs associations in the Middle East and North Africa region and they had diverse backgrounds in different aspects. We previously converted the PERSIAN FFQ used in the RaNCD cohort into NOVA groups; the results were as follows: unprocessed 59.5%, processed culinary 10.2%, processed 27.1%, and UPFs 3.0%; the same NOVA groups were used in this work. We found no significant association between UPF consumption and HTN incidence after adjusting the data in two different regression models (logistic regression and Cox proportional); the p-for trend was also insignificant. Several hypotheses can be made to justify not finding an association; firstly, the consumption of UPFs in our participants was extremely low (3.0%) compared to other studies; Martinez-Perez et al. reported the mean UPFs consumption in grams per day was 7.9% (7); Monge et al. and da Silva Scaranni et al. measured UPFs consumption percentages as total energy consumed, and found UPFs contributions were 29.8% and 25.2% respectively (8,9). Our results suggest that consuming UPFs in small quantities is not associated with HTN incidence. Secondly, we strictly adjusted covariates and tried to be cautious about categorizing controversial food items as UPFs. Previous studies had reported mixed results regarding UPFs/HTN association; some found a positive association. Mendonça et al., for example, used the Cox proportional regression model and found participants in the highest tertile of UPF consumption had adjusted HR (95%CI, p for trend) 1.21(1.06, 1.37, 0.004); some others used a wide range of statistical models including Poisson regression, linear regression, and t-test and found no significant associations (7,8,10). Fitting the data into two different regression models (logistic and Cox) helped us to have a clearer view of the association between HTN incidence and other covariates; when accounting for temporal aspects of HTN incidence (HRs) we found that females have 28% more chance to develop HTN compared to males; we also found that one year increase in age corresponds to 5% higher chance of HTN incidence; furthermore, we found that when other covariates are constant married and widowed participants had 114% and 130% respectively had higher chance of developing HTN. We also found that in our population, each unit increase in BMI corresponds to a 4% higher chance of HTN incidence; surprisingly, we found that one unit increase in WHR corresponds to a 654% higher chance of HTN incident, given the fact that difference in WHR among individuals is hugely lower than one unit it is better to say each 0.01 increase in WHR corresponds to 6.5% of rise in HTN incidence; because WHR is the best screening measure for CVD compared to other anthropometric indicators in Iranians, the implications of our findings become more noteworthy (11). Looking at familial aspects of HTN incidence, we found that a positive HTN history in first-degree relatives is associated with a 24% higher chance of HTN incidence. It is worth mentioning that calculated ORs were slightly higher than the mentioned HRs.

Our study, while comprehensive, did face some limitations. The PERSIAN FFQ, while not specifically designed for determining NOVA food groups, was manually determined by our team. We also encountered instances of missing data, and we chose to analyze the records with complete data. Despite our best efforts to adjust the model for all relevant confounders, there may be some unidentified ones. Lastly, longer follow-up periods could potentially reveal different results. These limitations, while present, do not diminish the robustness of our research. Our study has some substantial strengths as well. We are the first cohort study to assess the association of UPFs/HTN in the Middle East region. We bilaterally modified the main model into two different regression models, and we were able to calculate both ORs and HRs of confounders; additionally, we designed several sensitivity analysis scenarios to assess the integrity of the main model; furthermore, our cohort participants had a diverse background, and they did not belong to a specific group; diverse backgrounds of participants add to the generalization of our results.

In conclusion, our study, conducted using the logistic regression and Cox proportional hazard models, did not find a significant association between HTN Incidence and UPF consumption. However, we did find significant associations between HTN incidence and other covariates, including sex, age, marital status, BMI, WHR, FBS, and familial history of HTN. These findings, while not definitive, are significant and call for further studies with longer time frames and more participants to verify and expand upon our results.

## Data Availability

All data produced in the present study are available upon reasonable request to the authors

## Ethics Approval

Ethical approval for the study was obtained from the Kermanshah University of Medical Sciences Ethics Committee.

## Acknowledgments

We thank the participants of the RaNCD cohort for their cooperation and participation. We also want to thank all members of the RaNCD cohort for their unwavering support.

